# Social, demographic and behavioural determinants of SARS-CoV-2 infection: A case-control study carried out during mass community testing of asymptomatic individuals in South Wales, December 2020

**DOI:** 10.1101/2021.04.06.21253465

**Authors:** Daniel Rhys Thomas, Laia Homar Fina, James P. Adamson, Clare Sawyer, Angela Jones, Kelechi Nnoaham, Alicia Barrasa, A. Giri Shankar, Chris J. Williams

**Affiliations:** Communicable Disease Surveillance Centre, Public Health Wales, Cardiff, Wales, UK; UK Field Epidemiology Training Programme; Cwm Taf Morgannwg University Health Board, Abercynon, Rhondda Cynon Taf, Wales, UK; Health Protection Division, Public Health Wales, Cardiff, Wales, UK

**Author notes:** Twitter @DanielRhysThom1. Contributors: CW conceived the study. DRhT, LHF, JA and CS designed the study and recruited of subjects. DRhT, LHF, JA and ABB contributed to the analysis. DRhT drafted the manuscript with contributions from all authors. The study was funded through existing NHS Wales budgets with no external funding. No competing interests. Patient consent for publication was not required.

## Abstract

**Background:** Between 21 November and 22 December 2020, a SARS-CoV-2 community testing pilot took place in the South Wales Valleys. Lateral flow tests were offered to all people aged over 10 years living, studying or working in the area.

**Methods:** We conducted a case-control study in adults taking part in the pilot using an anonymous online questionnaire. Social, demographic and behavioural factors were compared in people with a positive test (cases) and a sample of negatives (controls). Population attributable fractions (PAF) were calculated for factors with significantly increased odds following multivariate analysis.

**Results:** A total of 199 cases and 2,621 controls were recruited by SMS (response rates: 27.1% and 37.6% respectively). Following adjustment, cases were more likely to work in the hospitality sector (aOR: 3.39, 95% CI: 1.43-8.03), social care (aOR: 2.63, 95% CI: 1.22-5.67) or healthcare (aOR: 2.31, 95% CI: 1.29-4.13), live with someone self-isolating due to contact with a case (aOR: 3.07, 95% CI: 2.03-4.62), visit a pub (aOR: 2.87, 95% CI: 1.11-7.37), and smoke or vape (aOR: 1.54, 95% CI: 1.02-2.32). In this community, and at this point in the epidemic, reducing transmission from a household contact who is self-isolating would have the biggest public health impact (PAF: 0.2).

**Conclusion:** Infection prevention and control should be strengthened to help reduce household transmission. As restrictions on social mixing are relaxed, hospitality venues will become of greater public health importance, and those working in this sector should be adequately protected. Smoking or vaping may be an important modifiable risk factor.

**What is already known on this subject?:** Certain populations are known to be at risk of severe COVID-19: Older people, males, people in minority ethnic groups, people with pre-existing chronic disease or disability, and people in certain public-facing occupations. However, limited information exists on the factors associated with acquiring SARS-CoV-2 in the community.

**What this study adds?:** This study provides an insight into the most important factors determining community transmission of SARS-CoV-2. We found that transmission within the household was the most important source of SARS-CoV-2 infection. Working in the hospitality sector, and visiting the pub were associated with infection but at the time of this study were relatively infrequent exposures. Smoking or vaping had a small but significant effect. Working in education, living with someone working in education, having caring responsibilities, attending a healthcare appointment and visiting a supermarket, restaurant, gym or leisure centre were not associated with infection. Whilst these findings relate to a specific community at a specific time in the course of the epidemic when social restrictions were in place, the information will be useful in supporting policy decisions. Mass testing exercises present an opportunity to conduct epidemiological studies to gather information to inform the local and national epidemic response.

## Introduction

There is growing evidence that certain population groups are more likely to be affected by severe COVID-19. These include: Older people, males, pregnant women, and people with pre-existing chronic disease or disability.^1-4^ People in certain minority ethnic groups and those in public-facing occupations are also disproportionally affected,^5-8^ but this is a combination of the risks of acquisition and progression to severe disease.

A proportion of SARS-CoV-2 infections will present as asymptomatic or mild infections, particularly in younger people, ^9,10^ so studies of risk factors for acquiring infection based on those hospitalised will be biased. Compared to evidence on risks of severe infection, limited information is available on the social, demographic and behavioural factors associated with transmission of SARS-CoV-2 infection in the community. Information gathered through the Test, Trace, Protect programme focuses on forwards contact tracing rather than factors associated with acquisition of infection.

A pilot mass testing exercise was initiated in South Wales. Whole borough testing took place Merthyr Tydfil (population approximately 60,000)^11^ between 21^st^ November and 20^th^ December 2020, and was extended to lower Cynon Valley in Rhondda Cynon Taf County Borough Council (an area of about 25,000 population covering five electoral wards) from 5^th^ to 22^nd^ December 2020. This was the second such initiative in the UK, after a pilot scheme in Liverpool,^12^ and the first in Wales. Testing was offered at community settings to asymptomatic people aged 11 and over living, working or studying in the two areas. Symptomatic people were asked to seek tests through other routes. A total of 47,619 lateral flow tests were carried out at 12 testing centres in Merthyr Tydfil and at eight testing centres in the Lower Cynon Valley. Of these, 1,135 (2.4%) were positive. People taking part were older than those in the catchment areas, and more tests (55%) were carried out in women.

Rates of confirmed Covid-19 in this relatively deprived, former industrial area of the South Wales Valleys, have been consistently high.^13^ This testing exercise presented an opportunity to conduct an epidemiological study to obtain information on factors associated with transmission in a high incidence setting, in order to inform the ongoing response.

## Methods

### Study design

Unmatched case-control. Target population was adults (18 years and over) living, working or studying in Merthyr Tydfil County Borough or electoral wards in the lower Cynon Valley, Rhondda Cynon Taf County Borough selected because they were areas of persistently high incidence. The study population was adults (18 years and over) attending community testing for at least one lateral flow test (LFT). Cases were defined as all people attending community testing pilot receiving a positive LFT result. Controls were a sample of those with a negative LFT result, with a planned case:control ratio of 1:3.

### Recruitment of cases and controls

Data on the results of LFT were de-duplicated to provide the first LFT for each person. These data contained the test result and the mobile phone number which was provided on registration when attending for testing. Rolling recruitment was carried out during the mass testing period. We contacted all cases and for each case, we randomly sampled 10 individuals who were tested on the same day but had a negative test result (controls).

### Data collection

A questionnaire was designed in the software tool *Smart Survey*.^14^ All newly tested individuals with a positive result (cases) and the sample of negatives (controls) were sent a SMS text message through the government portal texting service ‘*notify*.*gov*’^15^ asking them to complete an anonymous self-administered online questionnaire accessed via a hyperlink. To distinguish between cases and controls, a different link was sent to each group (See: text message in Appendix A, and questionnaire in Appendix B). We asked 37 questions on demographic and social factors, including: age, ethnicity, and occupation, area of residence, household structure, caring responsibilities, and social interactions in the previous 10 days.

### Analysis

Analysis was carried out using Stata v14.^16^ Response rates for cases and controls were calculated. The age distribution of cases responding was compared to all cases, and the age distribution of controls was compared with the sample selected for recruitment using Spearman Rank test.

We constructed a directed acyclic graph to inform the analysis. Having symptoms was excluded from the multivariable analysis as this considered not to be in the causal pathway. Also, being in contact with a known COVID-19 case was excluded from multivariable analysis, as this would underlie all other associated factors.

Variables were grouped into four categories: i. personal characteristics, ii. occupational exposures, iii. household exposures and iv. social exposures. Unmatched univariate analysis was carried out using Stata v14 to identify social and demographic factors associated with testing positive. Odds ratios with 95% confidence intervals were calculated for each exposure variable using logistic regression. Small area deprivation status was assigned to cases and controls using their area of residence. Deprivation quintiles were calculated based on the distribution of Welsh Index of Multiple Deprivation^16^ assigned to lower super output areas (LSOA) in Wales. Each participant was then classified into a deprivation quintile based on their LSOA of residence.

Multivariate analysis was carried out by logistic regression to take account of potential confounders or effect modifiers, identified *a priori* or in the univariate analysis. First, all exposures were adjusted for all other exposures within each of the four exposure categories i. to iv. Those variables that remained significant at p<0.05 were included in a further multivariate analysis to identify those factors most important in predicting risk of infection. Due to collinearity between ‘Place of work’ and ‘key worker’ fields, three new binary fields were created from the ‘key worker’ field: ‘Health and social care worker’, ‘Transport worker’ and ‘Public service worker’.

Lastly, to assess the public health significance of the exposures identified through multivariable analysis, we calculated population attributable fractions with 95% confidence intervals for those exposures that remained positively associated with testing positive after adjustment using *punafcc* post-estimation command in Stata.^17^ Adjusted odds ratios were plotted against population attributable fractions to investigate the relationship between personal risk and public health impact.

### Data privacy and information governance

We carried out this study to inform the ongoing epidemic response; and as such this activity is covered by Public Health Wales’ Establishment Order; and the Covid-19 privacy statement.^18^ Notify is a UK Government run platform which is a secure mass texting service. Notify is compliant with the Data Protection Act and any user data uploaded (e.g. phone numbers) are deleted after 7 days. Data which passes through the system is encrypted. Notify has been assessed and approved by the Cabinet Office Senior Information Risk Officer (SIRO). The SIRO checks this approval once a year. Notify is suitable for sending messages classified as ‘OFFICIAL’ or ‘OFFICIAL-SENSITIVE’ under the Government Security Classifications policy.

## Results

### Response

SMS messages were sent to 735 positives and 6,970 negatives aged 18 years or over and for whom we had a valid phone number for. A total of 199 cases and 2,621 controls were recruited, giving response rates of 27.1% and 37.6% respectively.

Cases had a similar age distribution to all people testing positive during the pilot (Spearman’s Rank Correlation, p=0.07), but although negative controls recruited had the same modal age group (50-59 years) as those selected to take part, older people were over-represented in the control group (Spearman’s Rank Correlation, p=0.01).

### Symptoms

Nearly all (99.6%) of people attending the testing pilot reported being asymptomatic at the point of test registration. However, at the time of questionnaire completion, 87 of 198 (44%) cases taking part in the study reported symptoms compatible with Covid-19 (loss of sense of smell/taste, a new ongoing cough, or a fever) indicating that a proportion of those testing positive were pre-symptomatic.

### Factors associated with a positive LFT

Cases were more likely to be in younger age groups (Table 1). Only small numbers of cases (<10) and controls (81) classified themselves as being in an ethnic group other than white-British or Irish. Cases were slightly more likely to be in a White – other ethnicity (odds ratio: 1.23), but this was not statistically significant. The majority of cases and controls lived in areas classified as within the three most deprived quintiles. Cases were slightly more likely to live in the most deprived areas and slightly less likely to live in the least deprived areas but this effect did not reach statistical significance (Table 1).

**Table 1.** Personal characteristics in people testing positive for SARS-CoV-2 (cases) and controls, with odds ratios. Factors significantly associated with being a case in bold.

|  |  | Cases |  | Controls |  | Univariate analysis |  |  | Multivariable analysis<sup>1</sup> |  |  |
| --- | --- | --- | --- | --- | --- | --- | --- | --- | --- | --- | --- |
|  |  | Exposed | % | Exposed | % | Odds ratio | 95% CI | p-value | Adjusted odds ratio | 95% CI | p-value |
| **Age group** |  | *n* = 199 |  | *n* = 2621 |  |  |  |  |  |  |  |
|  | 18-20 | 4 | 2.0 | 46 | 1.8 | 2.77 | 0.88 - 8.70 | 0.081 | 1.38 | 0.35 - 5.35 | 0.644 |
|  | **21-29** | **26** | **13.1** | **215** | **8.2** | **3.85** | **2.00 - 7.42** | **0.000** | **2.54** | **1.17 - 5.50** | **0.018** |
|  | **30-39** | **37** | **18.6** | **405** | **15.5** | **2.91** | **1.57 - 5.38** | **0.001** | 1.75 | 0.84 - 3.68 | 0.137 |
|  | **40-49** | **47** | **23.6** | **507** | **19.3** | **2.95** | **1.63 - 5.35** | **0.000** | **2.02** | **1.00 - 4.08** | **0.050** |
|  | **50-59** | **57** | **28.6** | **675** | **25.8** | **2.69** | **1.50 - 4.81** | **0.001** | 1.87 | 0.95 - 3.70 | 0.072 |
|  | 60-65 | 13 | 6.5 | 295 | 11.3 | 1.40 | 0.65 - 2.99 | 0.379 | 1.20 | 0.53 - 2.74 | 0.664 |
|  | Over 65 | 15 | 7.5 | 478 | 18.2 | *ref.* | - | - | *ref.* | - | - |
| **Ethnicity** |  | *n* = 199 |  | *n* = 2618 |  |  |  |  |  |  |  |
|  | White British or Irish | 192 | 96.5 | 2,537 | 96.5 | *ref.* | - | - | *ref.* | - | - |
|  | White other | 4 | 2.0 | 43 | 1.6 | 1.23 | 0.43 - 3.46 | 0.696 | 1.39 | 0.48 - 4.03 | 0.541 |
|  | Any other background | 3 | 1.5 | 38 | 1.4 | 1.04 | 0.32 - 3.41 | 0.944 | 1.07 | 0.25 - 4.66 | 0.927 |
| **Welsh deprivation quintiles** |  | *n* = 181 |  | *n* = 2406 |  |  |  |  |  |  |  |
|  | Most deprived | 55 | 30.4 | 616 | 25.6 | 1.21 | 0.66 - 2.19 | 0.531 | 1.05 | 0.56 - 1.96 | 0.872 |
|  | 2nd most deprived | 65 | 35.9 | 823 | 34.2 | 1.07 | 0.60 - 1.91 | 0.823 | 0.96 | 0.53 - 1.77 | 0.906 |
|  | 3rd most deprived | 34 | 18.8 | 588 | 24.4 | 0.78 | 0.41 - 1.47 | 0.444 | 0.79 | 0.41 - 1.52 | 0.484 |
|  | 4th most deprived | 12 | 6.6 | 176 | 7.3 | 0.92 | 0.42 - 2.02 | 0.841 | 0.81 | 0.36 - 1.80 | 0.606 |
|  | Least deprived | 15 | 8.3 | 203 | 8.4 | *ref.* | - | - | *ref.* | - | - |
| **Residence in catchment area** |  | *n* = 179 |  | *n* = 2380 |  |  |  |  |  |  |  |
|  | **Yes** | **124** | **69.3** | **1864** | **78.3** | **0.62** | **0.45 - 0.87** | **0.005** | 0.79 | 0.55 - 1.13 | 0.193 |
| **Smoke or vape** |  | *n* = 196 |  | *n* = 2619 |  |  |  |  |  |  |  |
|  | **Yes** | **44** | **22.4** | **422** | **16.1** | **1.51** | **1.06 - 2.14** | **0.022** | **1.47** | **1.00 - 2.15** | **0.048** |
| **Place of work** |  | *n* = 193 |  | *n* = 2,504 |  |  |  |  |  |  |  |
|  | Working from home or Not currently working | 51 | 26.4 | 1,053 | 42.1 | *ref.* | - | - | *ref.* | - | - |
|  | Factory/industrial setting | 13 | 6.7 | 174 | 6.9 | 1.54 | 0.82 - 2.90 | 0.177 | 1.38 | 0.70 - 2.74 | 0.350 |
|  | **Social care setting** | **11** | **5.7** | **74** | **3.0** | **3.07** | **1.53 - 6.14** | **0.002** | **2.60** | **1.25 - 5.39** | **0.010** |
|  | Education | 17 | 8.8 | 281 | 11.2 | 1.25 | 0.71 - 2.20 | 0.440 | 0.98 | 0.52 - 1.87 | 0.962 |
|  | **Healthcare setting** | **28** | **14.5** | **242** | **9.7** | **2.39** | **1.48 - 3.87** | **0.000** | **1.95** | **1.14 - 3.36** | **0.016** |
|  | **Hospitality** | **11** | **5.7** | **42** | **1.7** | **5.41** | **2.63 - 11.12** | **0.000** | **4.93** | **2.29 - 10.60** | **0.000** |
|  | Retail | 5 | 2.6 | 113 | 4.5 | 0.91 | 0.36 - 2.34 | 0.850 | 0.81 | 0.31 - 2.12 | 0.664 |
|  | **Office setting** | **28** | **14.5** | **245** | **9.8** | **2.36** | **1.46 - 3.82** | **0.000** | **2.13** | **1.24 - 3.66** | **0.006** |
|  | **Outside** | **6** | **3.1** | **50** | **2.0** | **2.48** | **1.02 - 6.05** | **0.046** | 2.28 | 0.91 - 5.72 | 0.080 |
|  | **In Prisons** | **1** | **0.5** | **1** | **0.0** | **20.65** | **1.27 - 334.82** | **0.033** | 12.25 | 0.72 - 209.58 | 0.084 |
|  | In homes/businesses/premises you are not resident in | 15 | 7.8 | 173 | 6.9 | 1.79 | 0.98 - 3.25 | 0.056 | 1.62 | 0.85 - 3.08 | 0.142 |
|  | Transport inc. deliveries | 1 | 0.5 | 22 | 0.9 | 0.94 | 0.12 - 7.10 | 0.951 | 0.87 | 0.11 - 6.72 | 0.895 |
|  | **Other** | **6** | **3.1** | **34** | **1.4** | **3.64** | **1.46 - 9.07** | **0.005** | 2.64 | 0.87 - 7.98 | 0.086 |
| **Key worker** |  | *n* = 197 |  | *n* = 2571 |  |  |  |  |  |  |  |
|  | Not a key worker or not currently working | 82 | 41.6 | 1269 | 49.4 | *ref.* | - | - | *ref.* | - | - |
|  | **Health and social care** | **45** | **22.8** | **406** | **15.8** | **1.72** | **1.17 - 2.51** | **0.005** | 1.26 | 0.82 - 1.93 | 0.286 |
|  | Public safety | 6 | 3.0 | 48 | 1.9 | 1.93 | 0.80 - 4.65 | 0.141 | 1.53 | 0.62 - 3.78 | 0.359 |
|  | Local and national government | 10 | 5.1 | 207 | 8.1 | 0.75 | 0.38 - 1.47 | 0.397 | 0.52 | 0.25 - 1.08 | 0.080 |
|  | Education and childcare | 19 | 9.6 | 321 | 12.5 | 0.91 | 0.55 - 1.53 | 0.738 | 0.62 | 0.35 - 1.11 | 0.107 |
|  | Food and necessary goods | 12 | 6.1 | 113 | 4.4 | 1.64 | 0.87 - 3.10 | 0.126 | 1.22 | 0.63 - 2.36 | 0.554 |
|  | **Transport** | **8** | **4.1** | **46** | **1.8** | **2.69** | **1.23 - 5.89** | **0.013** | 1.58 | 0.64 - 3.89 | 0.324 |
|  | Utilities, comms and financial services | 11 | 5.6 | 145 | 5.6 | 1.17 | 0.61 - 2.25 | 0.630 | 0.78 | 0.38 - 1.62 | 0.504 |
|  | **Public service worker** | **4** | **2.0** | **16** | **0.6** | **3.87** | **1.26 - 11.83** | **0.018** | **3.59** | **1.12 - 11.51** | **0.032** |

<sup>1</sup> Multivariable analysis adjusted for all other variables in table except 'key worker'. Multivariable analysis of 'key worker' was carried out by adjusting for all variables in the tables except 'Place of work'.

Most cases and controls were resident within the catchment, but cases were less likely to be resident inside the catchment area (OR: 0.62, 95% CI: 0.45-0.87). Twenty-two percent of cases reported smoking or vaping compared to 16% of controls (OR: 1.51, 95% CI: 1.06-2.14). Twenty-six percent of cases (51/193) were either not working or were working from home, as compared to 42% of controls. Compared to those not currently working or working from home, cases were more likely to work in a social care setting (OR: 3.07, 95% CI: 1.53-6.14), in a healthcare setting (OR: 2.39, 95% CI: 1.48-3.87), in hospitality (OR: 5.41, 95% CI: 2.63-11.12), in an office (OR: 2.36, 95% CI: 1.48-3.82), in prison (OR: 20.65, 95% CI: 1.27-334.82), or in an ‘other’ setting (OR: 3.64, 95% CI: 1.46-9.07). In those who worked, cases were less likely to work from home (OR: 0.43, 95% CI: 0.52-0.73)(Table 2).

**Table 2.**
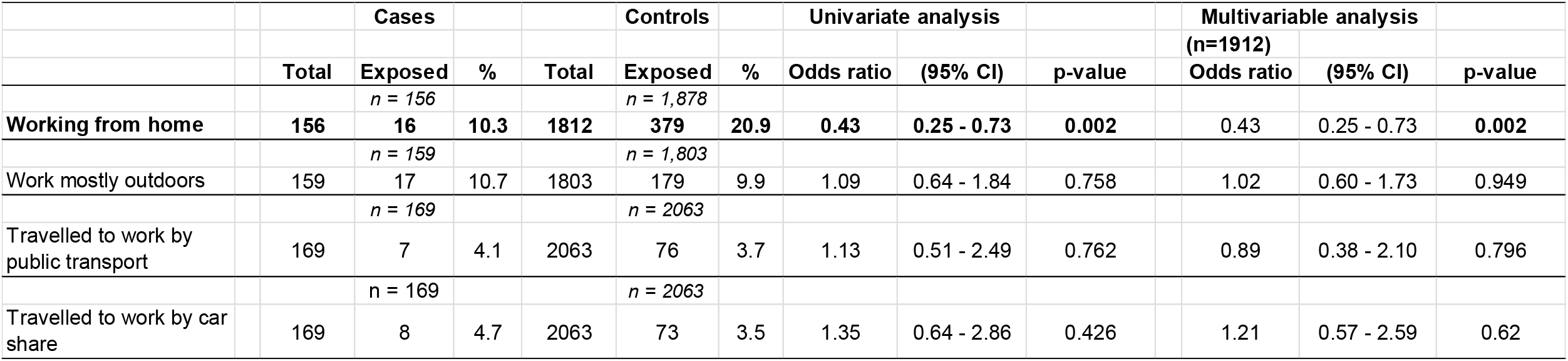
Occupational exposures in people who reported that they work. Factors significantly associated with being a case in bold. Multivariable analysis carried out by adjusting for all other variables in the table.

Univariate analysis of household exposures (Table 3) indicates that cases were more likely to live in larger households (odds ratio for living with 6 or more people: 4.43, 95% CI: 1.79-10.95, using living alone as a reference) were more likely to live with a child aged under eleven years (OR: 1.41, 95% CI: 1.01-1.97), were more likely to live with someone aged 23-59 years (OR: 1.60, 95% CI: 1.16-2.19) and were more likely to live with a healthcare worker (OR: 1.60, 95% CI: 1.08-2.37). Cases were less likely to live with someone aged 60 years or over (OR: 0.63, 95% CI: 0.44-0.90) or live with someone working in education (OR: 0.52, 95% CI: 0.27-0.99).

**Table 3.**
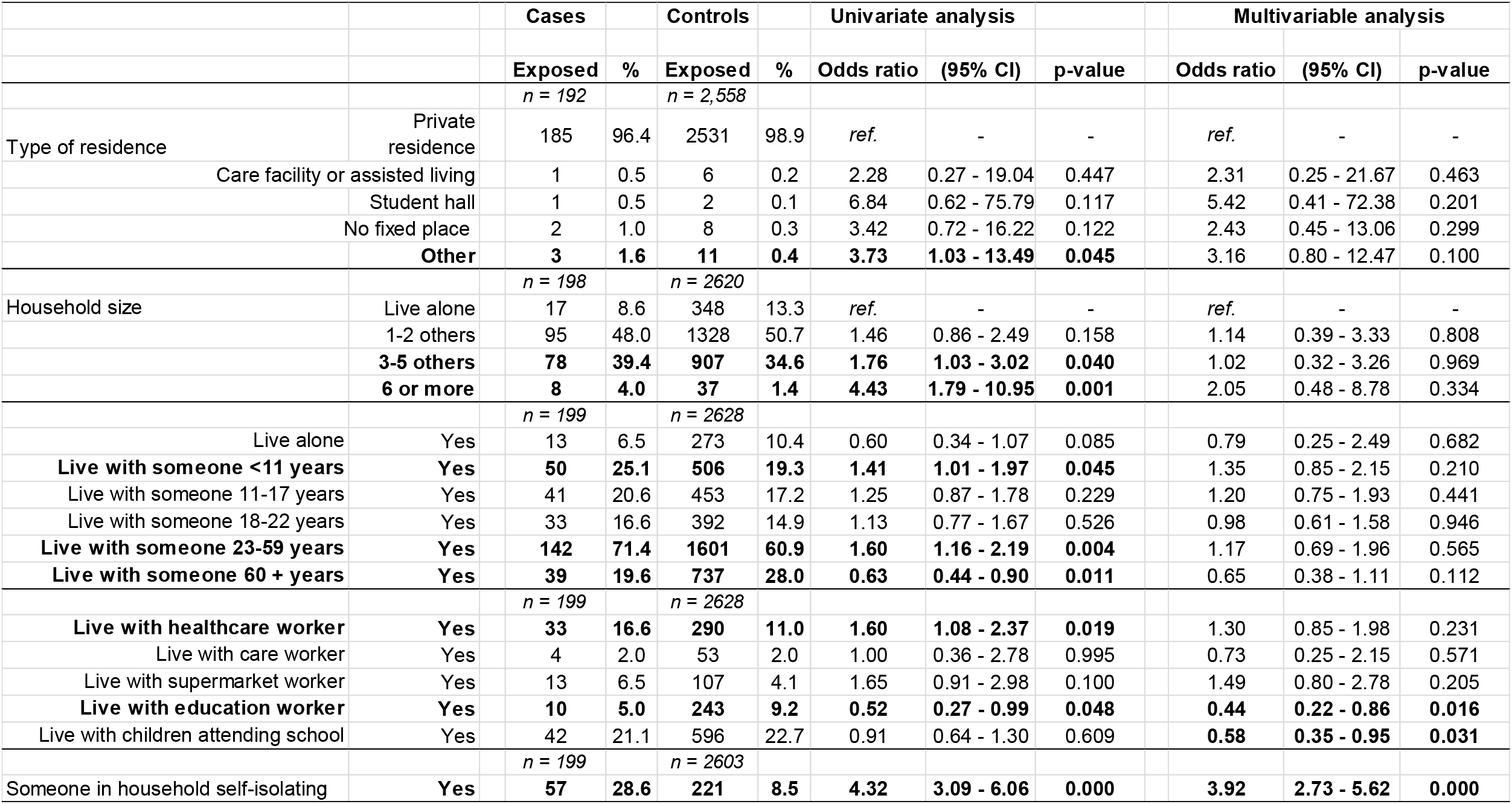
Household exposures in people testing positive for SARS-CoV-2 (cases) and controls, with odds ratios. Factors significantly associated with being a case in bold. Multivariable analysis carried out by adjusting for all other variables in the table.

Although only a small number of respondents visited a pub in the preceding 10 days (8 cases, 38 controls), this was significantly associated with infection (OR: 2.85) (Table 4). Cases were significantly less likely to have had household visitors, and were less likely to visit a shop or supermarket. Cases were not more likely to have caring responsibilities for someone outside their household. Cases were significantly less likely to have attended a face-to-face healthcare appointment in the preceding 10 days.

**Table 4.**
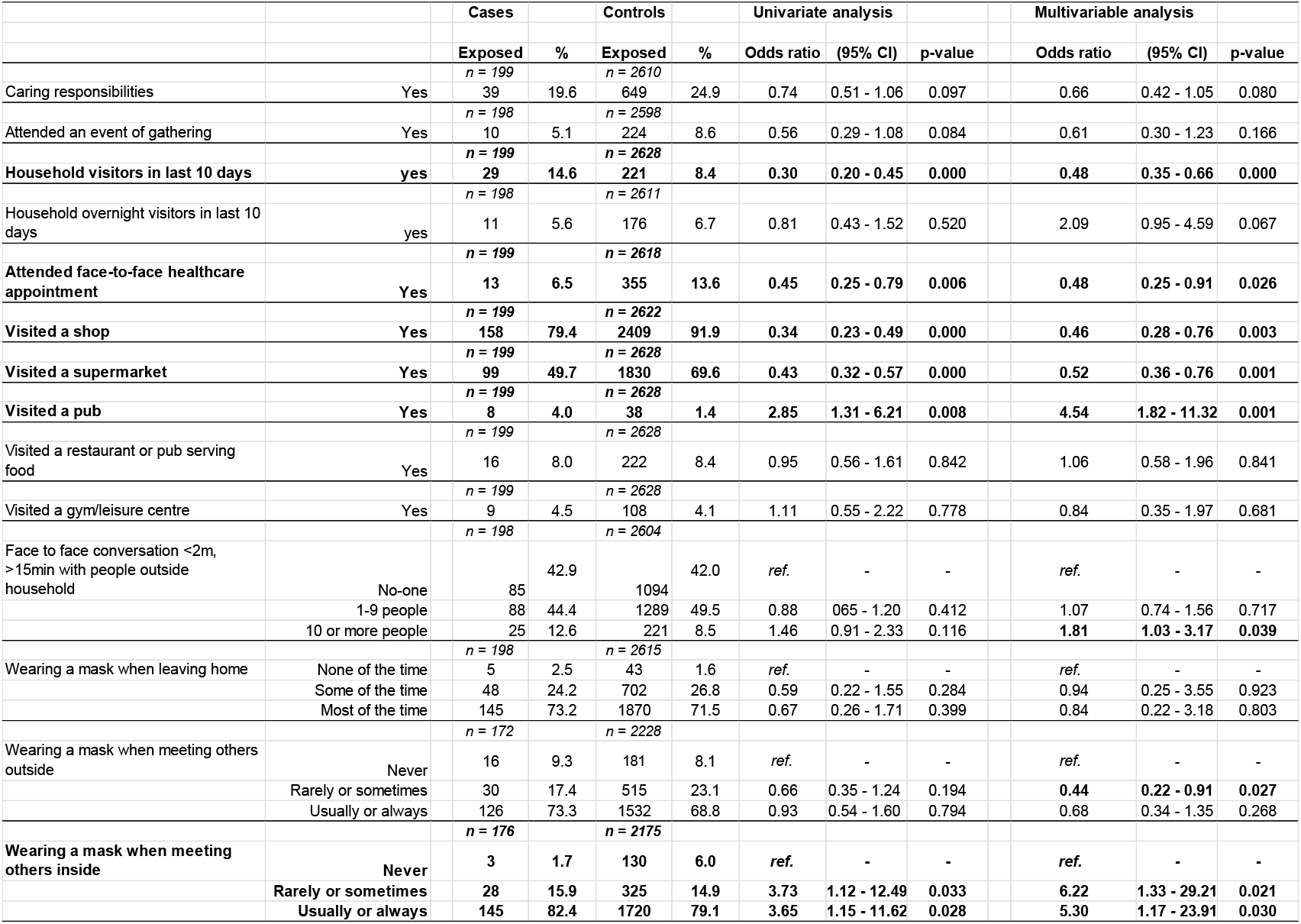
Social contact in people testing positive for SARS-CoV-2 (cases) and controls, with odds ratios. Factors significantly associated with being a case in bold. Multivariable analysis carried out by adjusting for all other variables in the table.

Cases were more likely than controls to report having been in contact with someone who has been told that they have a positive COVID-19 test in the last 10 days (odds ratio: 2.23, 95% confidence intervals: 1.63-3.05), and more likely to report someone in the household currently self-isolating because they had been in contact with someone with COVID-19 (odds ratio: 4.32, 95% confidence intervals: 3.09-6.06).

When asked about wearing face masks, most people (>70%) reported wearing a mask most of the time when leaving home. Cases reported being more likely to wear a face mask when meeting others inside. This remained significant after adjusting for all other social contact variables (Table 4).

Multivariate analysis identified working in the hospitality sector (pubs, bars, restaurants, hotels, betting shops) (aOR: 3.39, 95% CI: 1.43-8.03), working in a social care setting (aOR: 2.63, 95% CI: 1.22-5.67) working in a healthcare setting (aOR: 2.31, 95% CI: 1.29-4.13), living with someone who is self-isolating (aOR: 3.07, 95% CI: 2.03-4.62), visiting a pub in the preceding 10 days (aOR: 2.87, 95% CI: 1.11-7.37), and smoking or vaping (aOR: 1.54, 95% CI: 1.02-2.32) as the most important factors (Figure 1).

**Figure 1.**
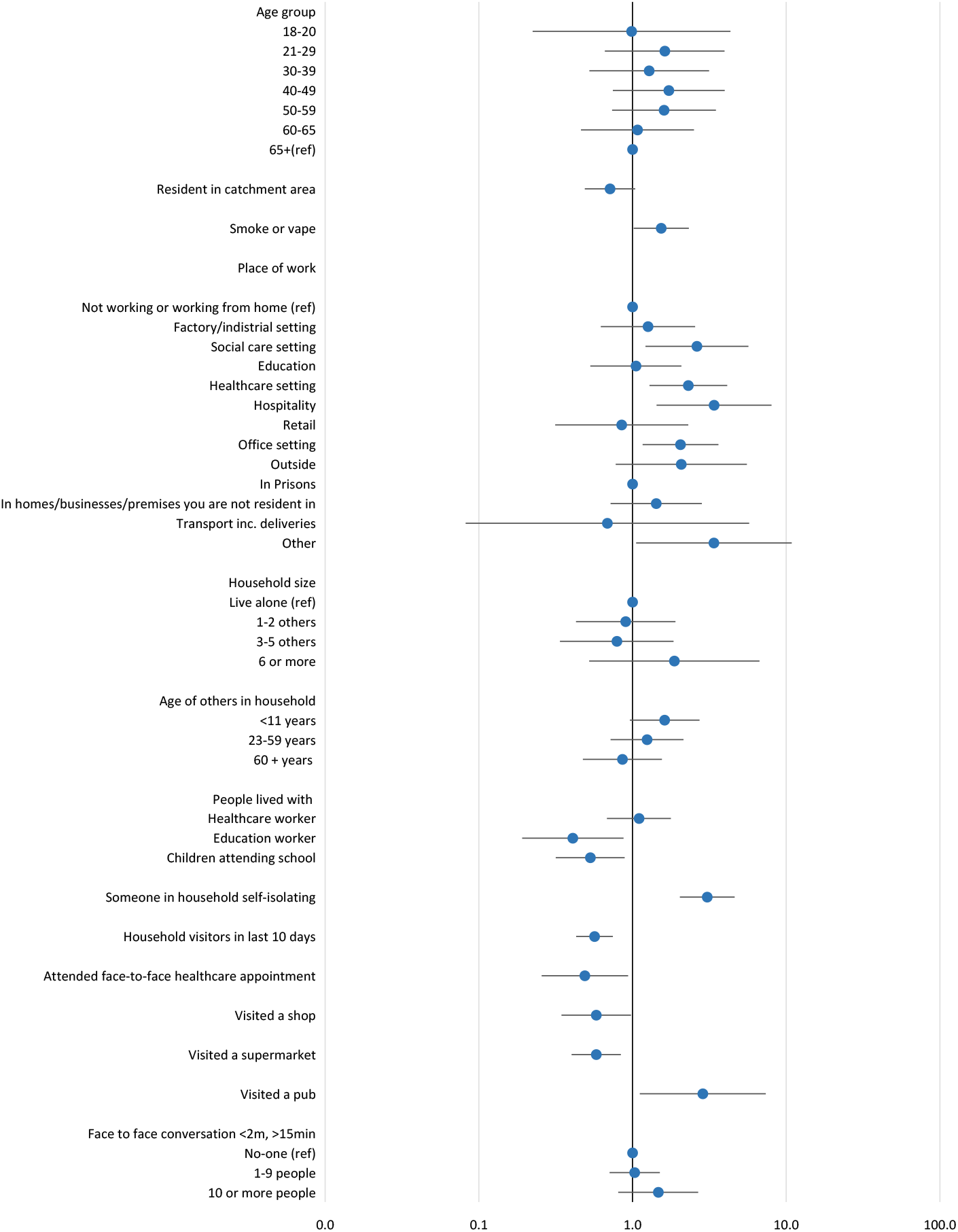
Forest plot showing adjusted odds ratios (aOR) for determinants of testing positive for SARS-CoV-2 in two areas of South Wales taking part in a community testing pilot, December 2020. aOR with 95% confidence intervals are given for those factors significant (p<0.05) in univariate analysis. Odds ratios greater than one represent an increased risk; odds ratios less than one represent a decreased risk. 95% confidence intervals not crossing one reflect that the odds ratio is statistically significant.

### Population attributable fractions

Population attributable fractions were 0.040 (95% CI: 0.020-0.059) for working in the hospitality sector, 0.033 (95% CI: 0.011-0.055) for working in a social care setting, 0.063 (95% CI: 0.024-0.100) for working in a healthcare setting, 0.204 (95% CI: 0.166-0.241) for living with someone who is self-isolating because they had been in contact with a confirmed case, 0.027 (95% CI: 0.015-0.040) for visiting a pub in the preceding 10 days, and 0.087 (95% CI: 0.021-0.149) for smoking or vaping (Figure 2).

**Figure 2.**
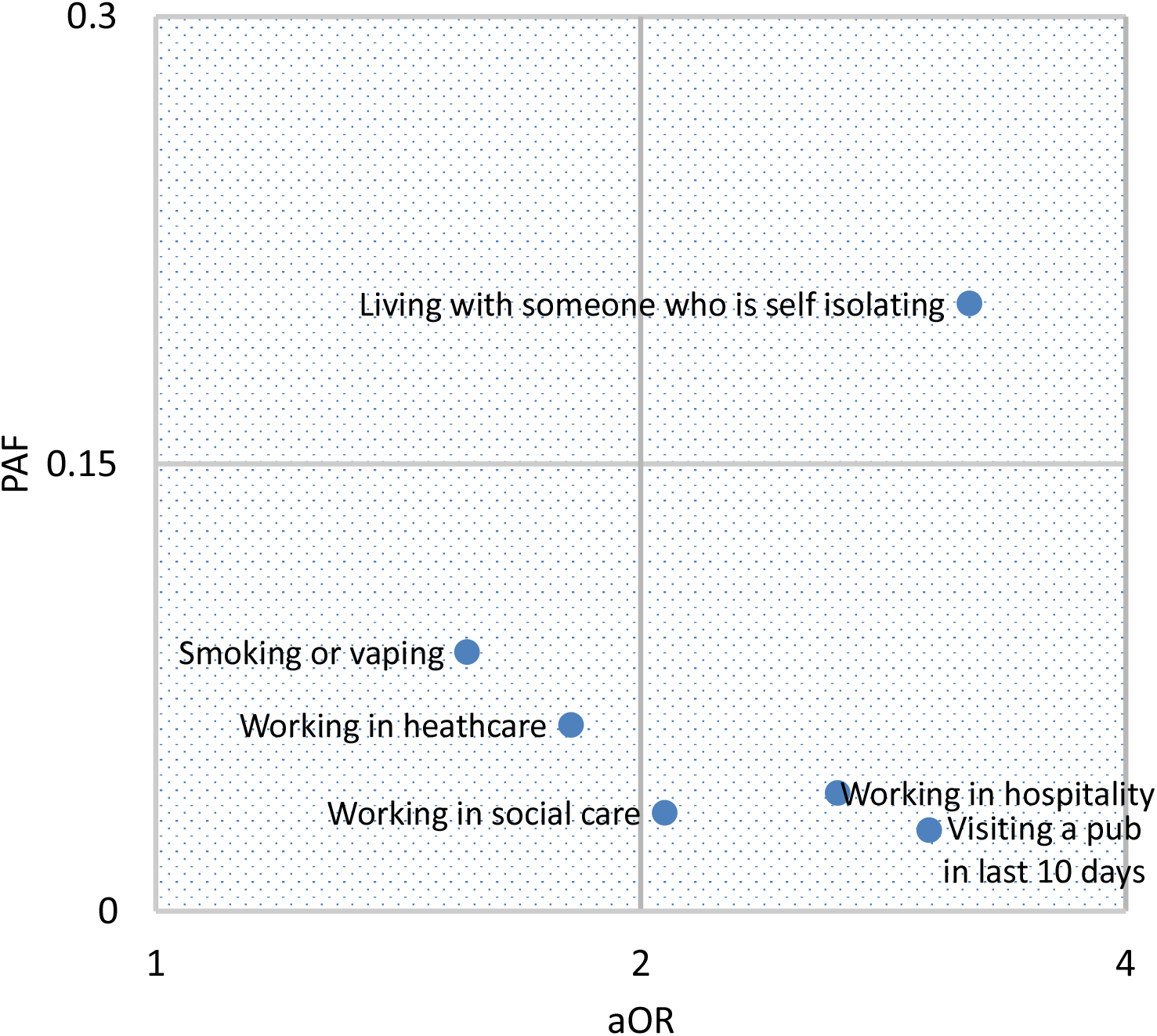
Relationship between personal risk, expressed as adjusted odds ratio (aOR) and public health impact expressed as population attributable fraction (PAF) for exposures associated with testing positive during the SARS-CoV-2 mass testing pilot in Merthyr Tydfil and lower Cynon Valley, 21 November to 20 December, 2020. aOR is plotted on a log scale.

## Discussion

This study provides insight into the most important factors determining community transmission of SARS-CoV-2. The study was carried out at the peak of the second wave of COVID-19 in the UK, and took place in localities which at the time had some of the highest rates of infection in the UK.

In this community, transmission within the household was an important source of SARS-CoV-2 infection. Household mixing is largely hidden, and may be perceived as lower risk than mixing with people from outside the home.^19^ Whilst media attention has focussed on adherence to restrictions affecting social contact outside the home, for example: travelling to exercise, attending work or going to school, transmission within households is being increasingly recognised as an important factor in the epidemiology of SARs-CoV-2.^20,21,22^ The former mining areas of the South Wales Valleys are characterised by close-knit communities, and have similarities with post-industrial towns in the North of England. One in five asymptomatic infections could have been prevented by avoiding contact with someone within the same household. Further work should be carried out to better understand the barriers to infection prevention and control within households, and how best to strengthen prevention and control advice, for example using online tools.^23,24^

Working in the hospitality sector, and visiting the pub were significant risks but at the time of this study were relatively infrequent exposures. The study took place before national ‘lockdown’ restrictions were introduced in Wales on 20 December, but were during a time when activity in the hospitality sector was restricted.^25,26^ As restrictions on social mixing are relaxed it is likely that exposure in hospitality venues will become of greater public health importance, and people working in this sector should be protected.

Smoking and vaping are potentially modifiable risk factors, and should be investigated further. Evidence for an association between smoking and COVID-19 has been mixed. Some researchers have suggested biological bases for an association. Others have suggested that it may relate to increased ‘hand to mouth’ contact. ^27,28^ Smoking confound other risk behaviours not measured in this study.

Of equal interest are the exposures that were not associated with infection. The policy to close schools and colleges has been debated, with concern that transmission risks are outweighed by the harms caused to children through lost education and socialisation.^29^ We found no evidence that education settings provided a significant risk of transmission to adults: Working in education, living with someone working in education, or living with school age children were not associated with testing positive.

The safety of supermarkets, restaurant, gyms and leisure centre has also been debated.^30^ Visiting these facilities did not appear to increase risk of infection.

Questions were asked about two specific non pharmaceutical interventions: Working from home and the wearing of face masks. Working from home was negatively associated with infection, and remains an important control measure. The results for mask wearing were unclear. In fact, in this study, people testing positive were more likely to report wearing a mask when meeting others inside. Qualitative methods could be used to investigate the behaviours associated with face mask use.

With so many associations investigated, it is always possible that some of our associations were chance findings. Moreover, statistically significant negative associations, such as living with an education worker, living with children who attend school, visiting a shop or supermarket, and attending a face-to-face health appointment may be the result of confounding by an another unknown factor. For example, people attending a face-to-face health appointment may be more likely to be in a clinically vulnerable group and therefore may be mixing less.

With a response rate of less than 40% it is possible that participants in our study were not representative of those people taking up the offer of testing. Moreover, it is likely that those accepting a test were not representative of people living in the catchment areas. Analysis by Cwm Taf Morgannwg University Health Board found that those taking up testing were older and were resident in less deprived areas of the catchment area.

The questionnaire was designed as a quick online questionnaire, taking participants around 5-10 minutes to complete, with participants recruited by SMS text message. The personal mobile phone number used to recruit was that given by participants at time of registering for testing at the community testing site, and the number which their lateral flow device test result was subsequently texted to. However, it is possible that some people were excluded from our survey as they did not have a valid mobile phone number of their own, or that their digital literacy level was not sufficient to use the link to our online questionnaire.

All exposures were self-reported. Although this was an anonymous study, all responses to questions about behaviour may be subject to social desirability bias, and should be interpreted with caution.

As an oversight, we did not include ‘gender’ on our questionnaire, preventing us from investigating the role of gender in our analysis. Another possible limitation in this study is choice of outcome measure, lateral flow test positivity. LFT is considered to be specific but not particularly sensitive.^31,32,33^ There will be some misclassification of cases and controls, but given the prevalence of SARS-CoV-2 in this setting, this is not considered to have had any significant impact on the findings.

The power of the case-control study was restricted by the number of lateral flow device positives, the frequency of certain determinants (for example there were only two people in our study reporting working in a prison setting) and our response rate. Factors such as working in a prison whilst no longer significant after adjusting for other variables, would warrant further investigation in future studies.

Mass testing as a control measure has proved controversial,^34,35^ but where it is undertaken, associated epidemiological studies can add to the knowledge about transmission risks. Combining this with calculation of attributable fractions helps to focus on the major drivers of transmission, in order to. produce evidence-based responses.

## Data Availability

Data may be made available on request, on a case by case basis.

